# The efficacy and safety of Favipiravir in treatment of COVID-19: A systematic review and meta-analysis of clinical trials

**DOI:** 10.1101/2021.02.14.21251693

**Authors:** Soheil Hassanipour, Morteza Arab-Zozani, Bahman Amani, Forough Heidarzad, Mohammad Fathalipour, Rudolph Martinez-de-Hoyo

**Affiliations:** Gastrointestinal and Liver Diseases Research Center, Guilan University of Medical Sciences, Rasht, Iran.; Social Determinants of Health Research Center, Birjand University of Medical sciences, Birjand, Iran.; Master of Health Technology Assessment, Faculty of Health, Tehran University of Medical Sciences, Tehran, Iran.; Department of Pharmacology and Toxicology, Faculty of Pharmacy, Hormozgan University of Medical Sciences, Bandar Abbas, Iran.; Medical Doctor, MSN Labs Americas

**Keywords:** COVID-19, SARS-CoV-2, Efficacy, Safety, Meta-analysis

## Abstract

The novel coronavirus outbreak began in late December 2019 and rapidly spread worldwide, critically impacting public health systems. A number of already approved and marketed drugs are being tested for repurposing, including Favipiravir. We aim to investigate the efficacy and safety of Favipiravir in treatment of COVID-19 patients through a systematic review and meta-analysis. This systematic review and meta-analysis were reported in accordance with the PRISMA statement. We registered the protocol in the PROSPERO (CRD42020180032). All clinical trials which addressed the safety and efficacy of Favipiravir in comparison to other control groups for treatment of patients with confirmed infection with SARS-CoV2 were included. We searched electronic databases including LitCovid hub/PubMed, Scopus, ISI web of Sciences, Cochrane, and Scientific Information Database up to 31 December 2020. We assessed the risk of bias of the included studies using Cochrane Collaboration criteria. All analyses were performed using the Comprehensive Meta-Analysis software version 2, and the risk ratio index was calculated. Egger and Begg test was used for assessing publication bias. Nine studies were included in our meta-analysis. The results of the meta-analysis revealed a significant clinical improvement in the Favipiravir group versus the control group during seven days after hospitalization (RR=1.24, 95% CI: 1.09-1.41; P=0.001). Viral clearance was more in 14 days after hospitalization in Favipiravir group than control group, but this finding marginally not significant (RR=1.11, 95% CI: 0.98-1.25; P=0.094). Requiring supplemental oxygen therapy in the Favipiravir group was 7% less than the control group, (RR=0.93, 95% CI: 0.67-1.28; P=0.664). Transferred to ICU and adverse events were not statistically different between two groups. The mortality rate in the Favipiravir group was approximately 30% less than the control group, but this finding not statistically significant. Favipiravir possibly exerted no significant beneficial effect in the term of mortality in the general group of patients with mild to moderate COVID-19. We should consider that perhaps the use of antiviral once the patient has symptoms is too late and this would explain their low efficacy in the clinical setting.

## Introduction

The novel coronavirus (SARS-CoV-2) outbreak began in late December 2019 and rapidly spread worldwide, critically impacting public health systems ^1^. As of January 5, 2021, the Johns Hopkins Coronavirus Resource Center has reported 85,783,178 confirmed Global Covid19 cases and a total of 1,855,872 worldwide deaths ^2^. The clinical characteristics of Coronavirus disease 2019 (COVID-19) include respiratory symptoms, fever, cough, dyspnea, and pneumonia. In the absence of any established pharmacological agents, supportive care remains the cornerstone of clinical management for COVID-19 ^3^.

As of October 22, 2020, remdesivir, an antiviral agent, is the only drug approved for treatment of COVID-19 ^4, 5^. An emergency use authorization (EUA) for convalescent plasma was announced on August 23, 2020 ^6^. The Food and Drug Administration (FDA) issued an EUA for bamlanivimab on November 9, 2020 ^7^. An EUA was issued for baricitinib on November 19, 2020 for use, in combination with remdesivir ^8^, and for casirivimab and imdevimab on November 21 ^9^. On December 11, 2020 the first vaccine (BNT-162b2 SARS-CoV-2 vaccine) was granted an EUA by the FDA and the same was accepted for a second vaccine (mRNA-1273 SARS-CoV-2 vaccine) on December 18, 2020 ^10^.

Numerous collaborative efforts to discover and evaluate effectiveness of antivirals, immunotherapy, monoclonal antibodies (at least 319 treatments under investigation), and 236 vaccines have rapidly emerged according to “The Milken Institute” that maintains a detailed COVID-19 Treatment and Vaccine Tracker of research and development progress ^11^. Due to the urgency of the situation, a number of already approved and marketed drugs are being tested for repurposing, including Favipiravir ^12^.

Favipiravir, also known as T-705, a purine nucleic acid analog, is one of the antiviral candidates considered in several clinical trials. It is a chemical used experimentally and was created by the Japanese company Toyama, a subsidiary of Fuji Film, as reported initially by Furuta in 2002 ^13^. In 2014, it was approved in Japan as a backup choice for resistant influenza infection and since then have been approved in several countries and is indicated for the treatment of patients with mild to moderate COVID-19 disease ^14^. Favipiravir is an RNA-dependent RNA polymerase inhibitor. It is activated in its phosphoribosylated form (favipiravir-RTP) in cells, inhibiting viral RNA polymerase activity^15^.

As of the 12th October 2020, there are 37 studies registered in the ClinicalTrials.gov database to evaluate the utility of this repurposed drug to fight against COVID-19 ^16^.

Even though multiple articles about favipiravir are readily available for download online, including some systematic reviews and meta-analyses conducted on only two RCTs at this time ^17–21^, the scientific community may find it challenging to get an overview regarding the safety and efficacy of this drug. Therefore, we aim to provide this systematic review and meta-analysis of Favipiravir. To do so, we assess all available completed clinical trials till December 2020^3^.

## Methods

### Protocol and registration

This systematic review and meta-analysis were reported in accordance with the Preferred Reporting Items for Systematic Reviews and Meta-Analyses (PRISMA) statement ^22^. We registered the protocol in the International Prospective Register of Systematic Reviews (PROSPERO) (CRD42020180032). Also, we published this protocol in the BMJ Open journal ^3^.

### Eligibility criteria

All clinical trials (study design) which addressed the safety and efficacy of Favipiravir (intervention) in comparison to other control groups (comparison) for treatment of patients with confirmed infection with SARS-CoV2 (population) were included. There were no restrictions concerning gender, age, ethnicity, blinding, follow-up, or publication status. All publications in English and Farsi were included. The investigated outcomes include clinical improvement, viral clearance, transferred to ICU, supplemental oxygen therapy, adverse events, and mortality. It should be noted that some of the outcomes mentioned in the protocol were not analyzed due to the lack of sufficient data in the final included articles. Articles with unavailable full text in English or Farsi languages or whose full text is not accessible were excluded from the study. The studies with insufficient or incomplete data were not being incorporated.

### Information sources and search strategy

Two independent reviewers (MA-Z and SH) searched electronic databases including LitCovid hub/PubMed ^23^, Scopus, ISI web of Sciences, Cochrane, and Scientific Information Database (SID) ^24^ using keywords combination (MeSH term and free term), such as “2019 nCoV” OR 2019nCoV OR “2019 novel coronavirus” OR COVID-19 OR “new coronavirus” OR “novel coronavirus” OR “SARS CoV-2” OR (Wuhan AND coronavirus) OR “SARS-CoV” OR “2019-nCoV” OR “SARS-CoV-2” and Favipiravir OR Avigan until the end of 2020. We also searched two preprint databases including MedRxiv and Research Square and, the reference lists of all included studies, reviews, and clinical trial registries, for an ongoing clinical trial (see Supplementary file 1 for the final proposed PubMed search strategy).

### Study records

Once the records have been imported to EndNote X7 software and all duplicates have been removed, two reviewers (SH and BA) independently screened titles, abstracts, and full-texts of included studies based on predefined eligibility criteria to identify studies concerning the safety and efficacy of Favipiravir among patients with COVID-19. All potentials discrepancies were resolved upon consultation with a third reviewer (MA-Z).

### Data extraction and data items

Two reviewers (SH and BA) independently extracted data from included studies, using a pre-piloted data extraction form. We piloted this form using at least three examples of included studies and if there is a 90% and above agreement, it is approved. The data extraction form includes the following items; authors name, year of the publication, study design, study sample, country of origin, mean age of participants, gender, the severity of diseases, comorbidities, type of intervention and dose, control group, follow up, randomization, blinding, allocation concealment, primary and secondary outcomes, and adverse events ^3, 25^. All potentials discrepancies were resolved by consultation with a third reviewer (RM).

### Risk of bias in individual studies

Two reviewers (MF and FH) independently assessed the risk of bias among the included studies. We assessed the risk of bias of the included studies using Cochrane Collaboration criteria, including seven items of selection bias (random sequence generation and allocation concealment), performance bias, detection bias, attrition bias, reporting bias, and other forms of bias ^3, 25^. Any discrepancies were resolved upon consultation with a third reviewer (MA-Z).

### Statistical analysis

All analyses were performed using the Comprehensive Meta-Analysis (CMA; Borenstein, Hedges, Higgins, and Rothstein) software version 2, and the risk ratio (RR) index was calculated. CMA software has the ability to combine different indices and to combine the effect of sample size and the difference of the index being compared ^26^. We used the I^2^ statistics and Cochran test (with significantly less than 0.1) to assess the heterogeneity of the included studies ^27^. In cases where there was heterogeneity, we performed the random-effect model. We also used a subgroup analysis based on follow up days for clinical improvement and viral clearance. Egger and Begg test was used for assessing publication bias.

## Results

### Description of search

We identified a total of 1340 records after searching the databases. After the removal of 431 duplicate records, the title and abstracts of 909 records were screened. Eight hundred eighty-five records were excluded after title and abstracts screening, and 24 records were assessed for full-text screening. A total of 15 records were excluded based on eligibility criteria. Finally, nine studies were included in our meta-analysis ^28–36^ (Fig 1).

**Figure 1:**
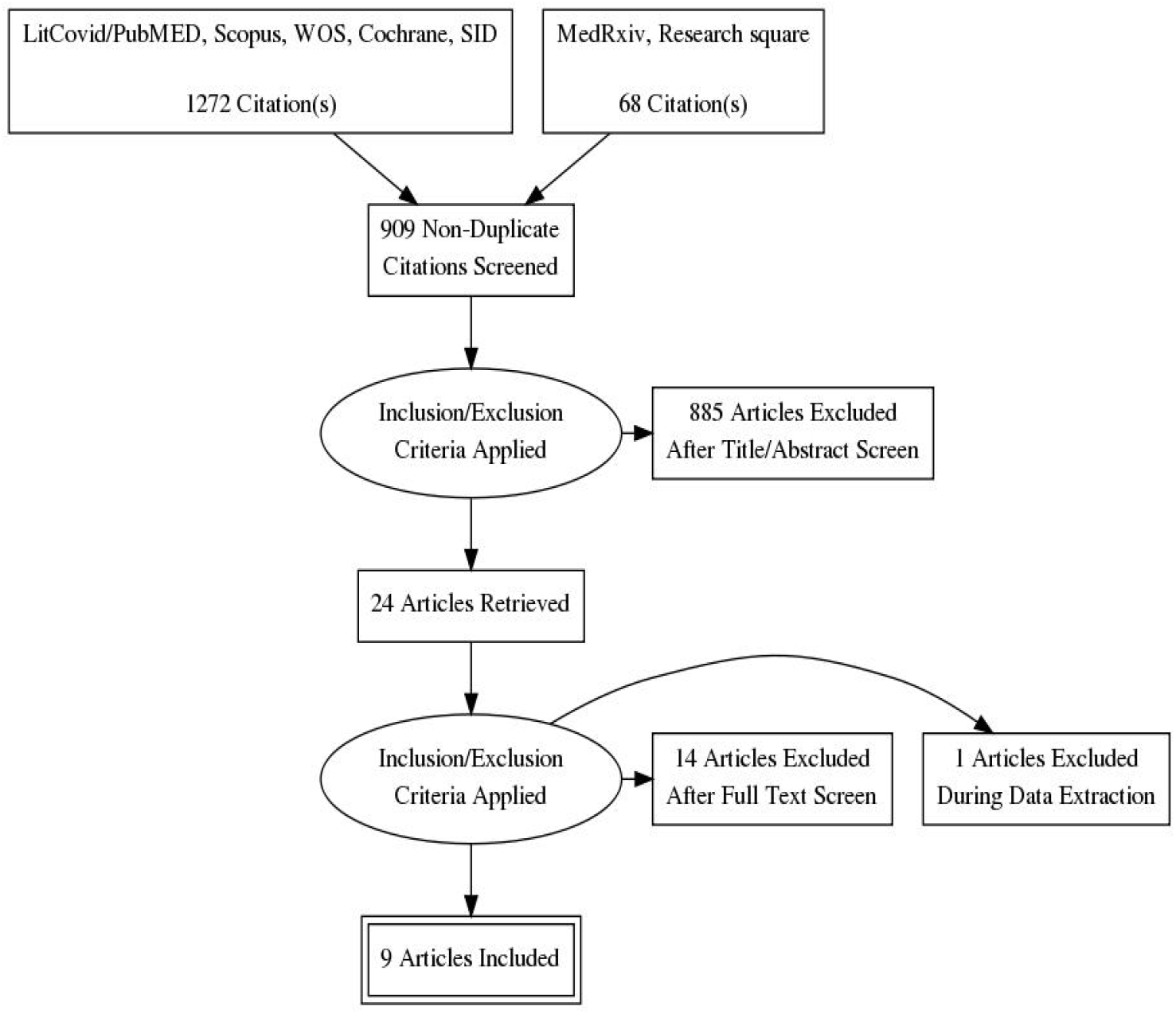
Search process and study flow diagram

### Characteristics of the included studies

Nine studies encompassing 827 patients were included. According to the geographical area, four studies were conducted in China (44.4%), and Russia, Oman, Egypt, and Japan also had an article. Only one study was nonrandomized. The minimum follow-up time was ten days, and the maximum was 30 days. The doses of Favipiravir and control drugs in each study were different. All studies registered in clinical trial registries. The summary characteristics of the included studies have been summarized in Table 1.

**Table 1:** summary characteristics of the included studies

| Authors/ year | ClinicalTrials Identifier | Publication status* | journal | Country | Study design | Age | Male | Intervention /sample size | Comparison/ sample size | Follow up |
| --- | --- | --- | --- | --- | --- | --- | --- | --- | --- | --- |
| Cai et al/ 2020 | ChiCTR2000029600 | In press | Engineering | China | Open-label, nonrandomized, before-after controlled study | 16–75 | 40% in FVP group/ 46.7% in LPV/RTV group | Oral FPV (Day 1: 1600 mg twice daily; Days 2–14: 600 mg twice daily) plus interferon (IFN)-a by aerosol inhalation (5 million U twice daily)/ 35 | LPV/RTV (Days 1–14: 400 mg/100 mg twice daily) plus IFN-a by aerosol inhalation (5 million U twice daily)/ 45 | Day 14 |
| Chen et al/ 2020 | ChiCTR2000030254 | Preprint | MedRxiv | China | prospective, randomized, controlled, open-label multicenter trial | 18 years or older | 50.8% in FVP group/ 42.5% in Arbidol group | FPV (1600mg*2/first day followed by 600mg*2/day) for 10 days/ 116 | Umifenovir (Arbidol) (200mg*3/day)/ 120 | Day 10 |
| Dabbous et al/ 2020 | NCT04349241 | Preprint | Research square | Egypt | Computer based randomized controlled interventional clinical trial phase 3 | 18-80 | 50% in each groups | FPV 3200mg at day1 followed by 600mg twice (day2-day10)/ 50 | HCQ 800mg at day1 followed by 200mg twice (day2- 10) and oral oseltamivir 75mg/12hour/day for 10 days/ 50 | Day 30 |
| Doi et al/ 2020 | jRCTs041190120 | In press | Antimicrobial agents and chemotherapy | Japan | prospective, randomized, open -label, multicenter trial | 16 years or older | 52.3% in early group, 705.% in late group | Early FPV: Favipiravir was dosed at 1,800 mg orally at least four hours apart on the first day, followed by 800<br>278 mg orally twice a day, for a total of up to 19 doses over 10 days/ 36 | Late FVP: Favipiravir was dosed at 1,800 mg orally at least four hours apart on the first day, followed by 800/ 33 | Day 28 |
| Ivashchenko et al/ 2020 | NCT04434248 | In press | Clinical infectious | Russia | Adaptive, multicenter, open label, | 18 years or older | NR | AVIFAVIR 1600 mg BID on Day 1 followed by 600 mg BID on Days 2-14 | Standard of care of Russian guidelines for treatment of | Day 29 |
|  |  |  | disease |  | randomized,<br>phase 2 and 3<br>clinical trial |  |  | (1600/600 mg)/ 20 | COVID-19/ 20 |  |
|  |  |  |  |  |  |  |  | AVIFAVIR 1800 mg BID<br>on Day 1 followed by 800<br>mg BID on Days 2-14<br>(1800/800 mg)/ 20 |  |  |
| Khamis et al/<br>2021 | NCT04385095 | Published<br>online | International Journal of<br>Infectious Diseases | Oman | Open label<br>randomized<br>controlled study | 18-75 | 64% in FVP<br>group/ 53% in<br>SOC group | FPV 1600 mg on day 1<br>followed by 600 mg twice<br>a day for a maximum of<br>10 days, and interferon<br>beta-1b at a dose of 8<br>million IU (0.25 mg)<br>twice a day was given for<br>5 days through a vibrating<br>mesh aerogen nebulizer/<br>44 | Standard of care of Oman<br>guidelines for treatment of<br>COVID-19: HCQ 400 mg<br>twice per day on day 1, then<br>200mg twice per day for 7<br>days/ 45 | Day 14 |
| Lou et al/<br>2020 | ChiCTR200002<br>9544 | Published | European Journal of<br>Pharmaceutical Sciences | China | Exploratory<br>single center,<br>open-label,<br>randomized,<br>controlled trial | Mean: 58,<br>53.5 and<br>46.6 for<br>FAV,<br>Baloxavir<br>and<br>control<br>group | 77% in FVP<br>group/ 70%<br>in other<br>groups | FAV group: 1600 mg or<br>2200mg orally, followed<br>by 600 mg each time,<br>three times a day, and the<br>duration of administration<br>was not more than 14<br>days/ 9 | Baloxavirmarboxil group:<br>80 mg once a day orally on<br>Day 1 and Day 4; for<br>patients who are still<br>positive in virological test,<br>they can be given again on<br>Day 7/ 10<br>Control group: LPV/RTV<br>(400 mg/100 mg, bid, po.)<br>or darunavir/cobicistat (800<br>mg/150 mg, qd, po.) and<br>arbidol (200 mg, tid, po.)/<br>10 | Day 14 |
| Udwadia et al/<br>2020 | CTRI/2020/05/0<br>25114 | Published | International Journal of<br>Infectious Diseases | India | Randomized,<br>open-label,<br>parallel-arm,<br>multicenter, | 18-75 | 70.8% in FVP<br>group/ 76%<br>in control<br>group | Oral favipiravir (1800mg<br>BID loading dose on day<br>1; 800 mg BID<br>maintenance dose<br>thereafter) plus standard | Standard supportive care<br>alone that included<br>antipyretics, cough<br>suppressants, antibiotics, | Day 14 |
|  |  |  |  |  | phase 3 study |  |  | supportive care for up to a maximum of 14 days/ 70 | and vita-mins/ 68 |  |
| Zhao et al/ 2021 | ChiCTR2000030894 and NCT04310228 | Published online | Biomedicine & Pharmacotherapy | China | Multicenter, randomized trial | 18 years or older | 71.4% in FVP group/ 60% in Tocilizumab group | FAV group: 1600mg, twice a day on the first day, and 600mg, twice a day from the second day to the seventh day, orally/ 7 | Combination group (FAV+ tocilizumab)/ 14<br>Tocilizumab group: first dose was 4–8mg/kg (recommended 400mg) and added to 100mL 0.9 % normal saline/ 5 |  |

NR: Not Reported, FPV: Favipiravir, LPV: Lopinavir, RTV: Ritonavir, HCQ: hydroxychloroquine
\* The status of manuscript in time of screening

### Risk of bias in individual studies

Eight (88.8%) studies described the random sequence generation. Six studies (66.6%) described the allocation concealment in an acceptable manner. None of the studies reported acceptable blinding for participants and personnel. Only one study (11.1%) reported blinding of outcome assessment. The risk of bias summary and risk of bias graph is reported in Supplementary files 2 and 3.

### The results of the meta-analysis

#### Clinical improvement

Among the included studies, six studies assessed clinical improvement during 14 days after hospitalization, and five studies were assessed during seven days after hospitalization. The results of the meta-analysis revealed a significant clinical improvement in the Favipiravir group versus the control group during seven days after hospitalization (RR=1.24, 95% CI: 1.09-1.41; P=0.001, I2= 0.0%, P=0.939). On the other hand, in 14 days after hospitalization, clinical improvement was 10% higher in the Favipiravir group, but this finding not statistically significant (RR=1.10, 95% CI: 0.97-1.25; P=0.108, I2= 34.5%, P=0.177) (Fig 2).

**Figure 2:**
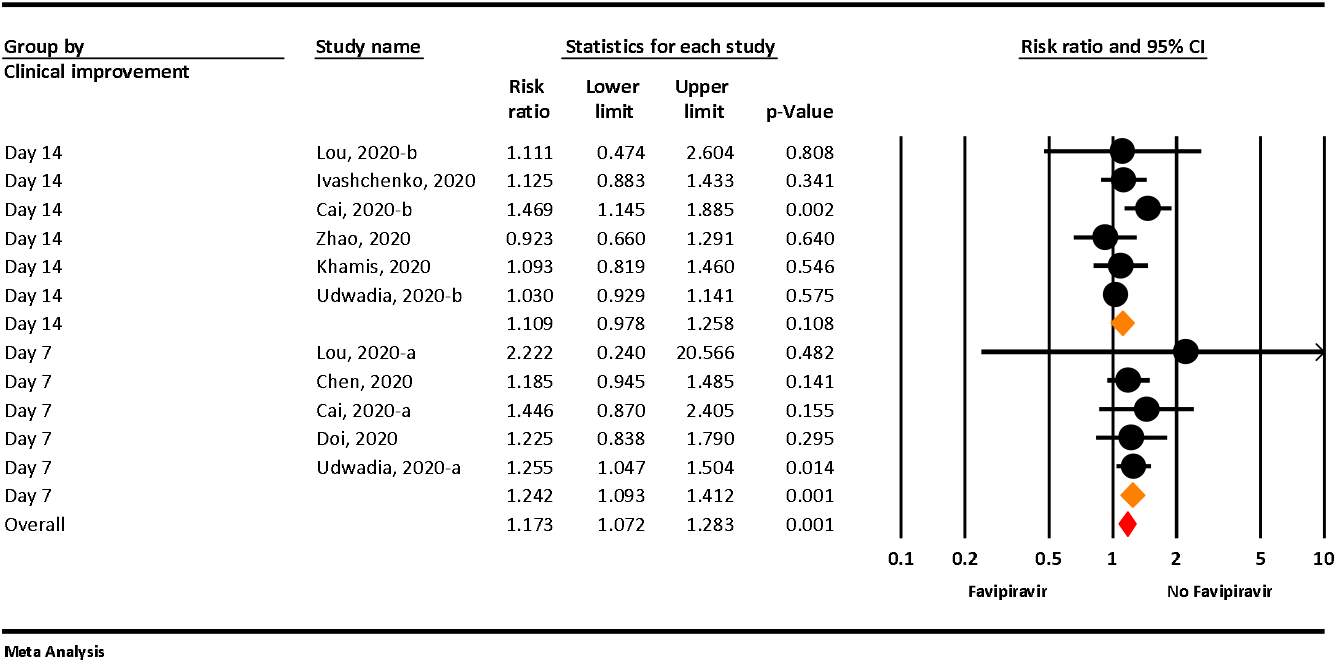
The meta-analysis of clinical improvement of Favipiravir on COVID-19 patients

#### Viral clearance

The result of meta-analysis show that, viral clearance was more in 14 days after hospitalization in Favipiravir group than control group, but this finding marginally not significant (RR=1.11, 95% CI: 0.98-1.25; P=0.094, I2= 42.9%, P=0.112). Viral clearance in 7 and 10 days after hospitalization not statistically different between two groups (RR=1.07, 95% CI: 0.83-1.39; P=0.580, I2= 62.1%, P=0.022 for 7 days and RR=1.02, 95% CI: 0.92-1.13; P=0.648, I2= 0.0%, P=0.846 for 10 days) (Fig 3).

**Figure 3:**
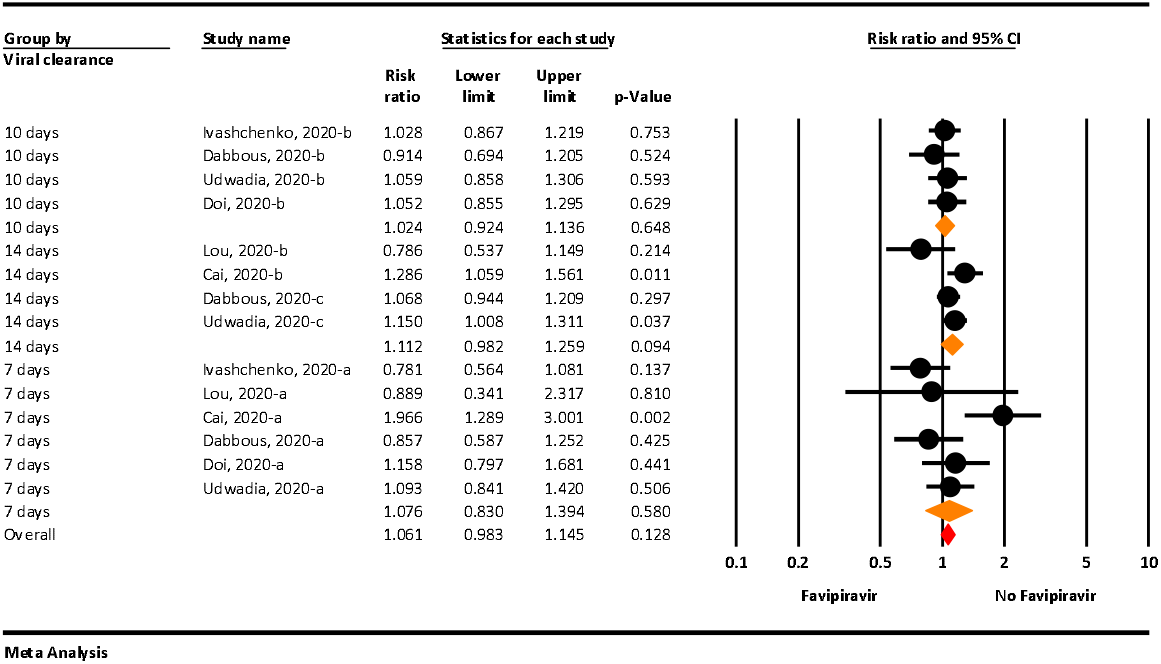
The meta-analysis of viral clearance of Favipiravir on COVID-19 patients

#### Requiring supplemental oxygen therapy

Based on the meta-analysis, requiring supplemental oxygen therapy in the Favipiravir group was 7% less than the control group, but this finding not statistically significant(RR=0.93, 95% CI: 0.67-1.28; P=0.664, I2= 0.0%, P=0.950) (Fig 4).

**Figure 4:**
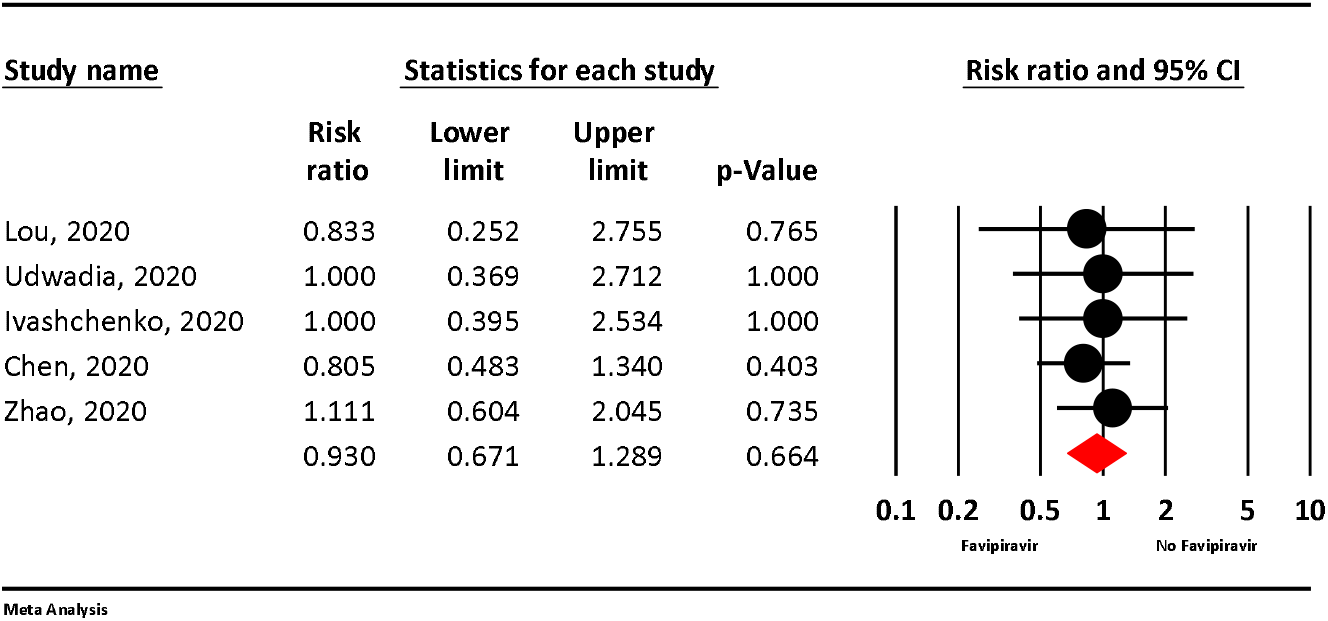
The meta-analysis of requiring supplemental oxygen therapy of Favipiravir on COVID-19 patients

#### Adverse Events

Meta-analysis comparing adverse events between the Favipiravir and the control groups showed lesser odds for adverse effects in the Favipiravir arm but of no statistical significance (RR=0.77, 95% CI: 0.34-1.70; P=0.524, I2= 85.4%, P<0.001) (Fig 5).

**Figure 5:**
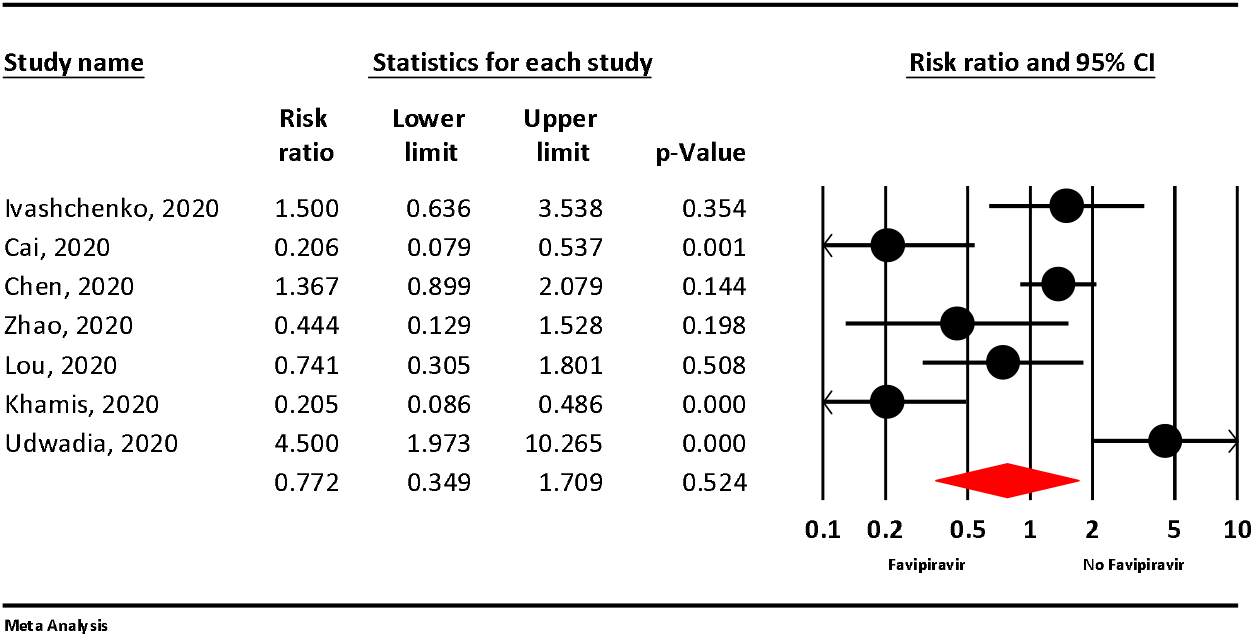
The meta-analysis of adverse events of Favipiravir on COVID-19 patients

#### Transferred to ICU

Based on meta-analysis, transferred to ICU not statistically different between two groups (RR=1.13, 95% CI: 0.49-2.59; P=0.759, I2= 0.0%, P=0.525) (Fig 6).

**Figure 6:**
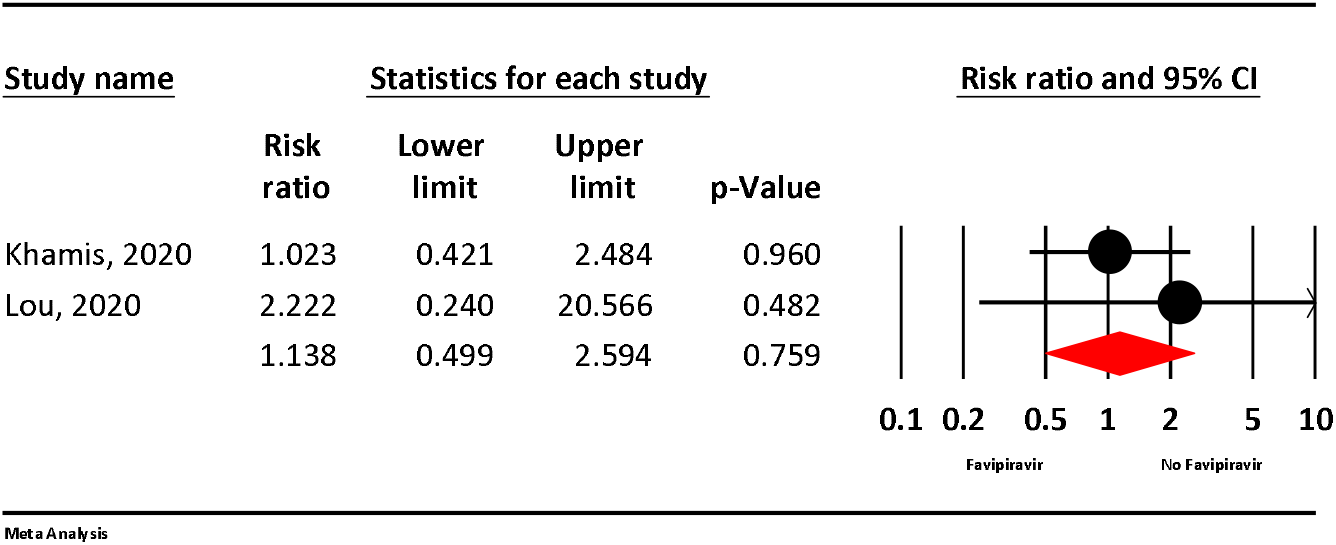
The meta-analysis of transferred to ICU of Favipiravir on COVID-19 patients

#### Mortality

Based on the meta-analysis, the mortality rate in the Favipiravir group was approximately 30 % less than the control group, but this finding not statistically significant (RR=0.70, 95% CI: 0.26-1.28; P=0.664, I2= 0.0%, P=0.950) (Fig 7)

**Figure 7:**
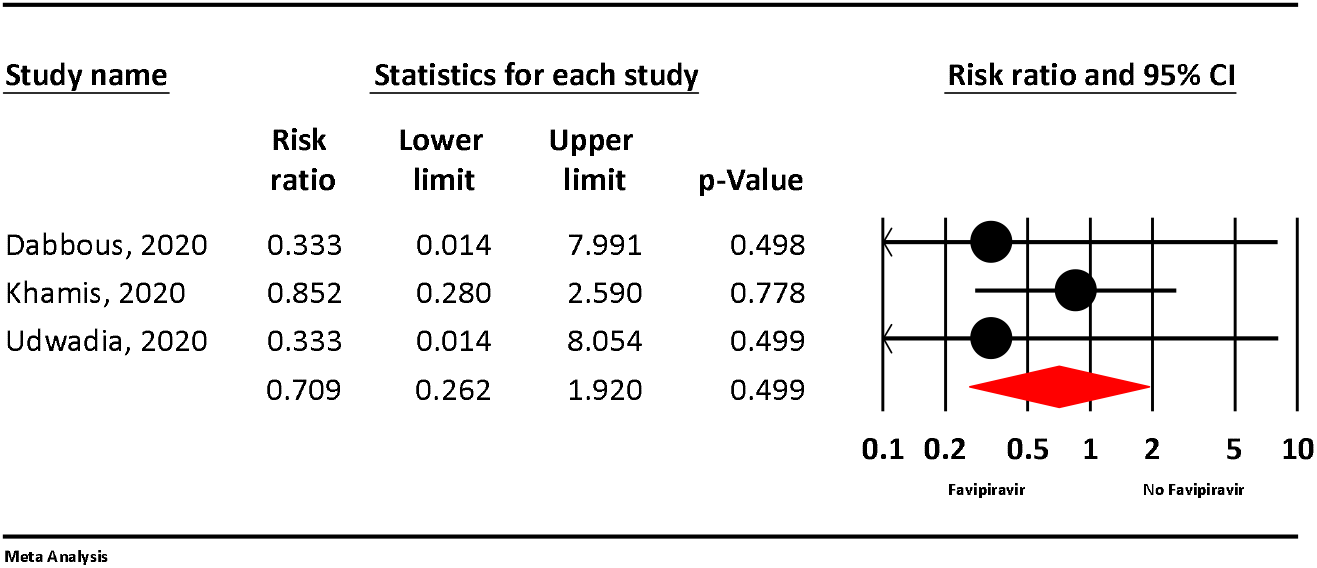
The meta-analysis of mortality of Favipiravir on COVID-19 patients

#### Publication bias

Publication bias was not observed among the included studies according to the results of the Egger (P=0.944) and Begg test (P=0.956).

## Discussion

The COVID-19 causing SARS-CoV-2, an acute respiratory disease, is spreading rapidly and has led to a pandemic with devastating effects within a few months of November 2019 ^37^.The number of infected cases, as well as the mortality rate associated with the virus, has astronomically raised around the world. The main challenge of COVID-19 is the lack of approved pharmacotherapy and vaccination, as well as the absence of evidence for reliable treatment options ^37^. Although various agents are undergoing clinical trials, the urgency of the situation has made scientists repurpose the antiviral agents.

Favipiravir, as a ribonucleotide analog and selective inhibitor of the viral RNA polymerase enzyme, can cause widespread antiviral activity against RNA-carrying viruses, thereby preventing replication and transcription of the viral genome ^38^. It has been approved for the treatment of new influenza viruses in Japan and China ^39^. It has also been shown to be effective against Ebola and RNA viruses caused by viral hemorrhagic fever ^30^.

However, none of the society and organizational guidelines (IDSA guidelines, World Health Organization guidelines, National Institutes of Health guidelines) recommend using favipiravir in the management of COVID-19, given the varying results of existing clinical trials data ^39^. Moreover, this drug revealed controversial results in different clinical trials conducted on COVID-19. Therefore, we decided to investigate the safety and efficacy of favipiravir in the treatment of COVID-19. Our meta-analysis was carried out on nine eligible studies with 827 patients.

The obtained results demonstrated the clinical improvement after seven and 14 days of hospitalization was more remarkable in patients taking favipiravir than those receiving other drugs. Another meta-analysis conducted by Shrestha et al. demonstrated that clinical improvement was observed on both the seventh and 14^th^ day of treatment ^40^. Udwadia et al. reported the time of clinical improvement was significantly faster in patients in the favipiravir group than those who are not ^39^.

The viral clearance after 14 days of hospitalization among patients taking favipiravir was more than those taking other drugs. However, this difference was not statistically significant after seven and ten days, which could be related to inappropriate dose and duration of treatment with favipiravir ^41^. In another meta-analysis by Shrestha et al., it was stated that viral clearance on seventh and 14^th^ was not significant between the favipiravir and control groups ^40^.

This difference between the results of our analysis and the mentioned meta-analysis might be due to an insufficient number of studies and a small sample size in the Shrestha et al. meta-analysis. Adolfo Pérez-García et al. reported that a randomized study on 80 patients with mild COVID-19 showed favipiravir group reduced virus clearance time by 50% compared to Lopinavir/Ritonavir group ^38^.

Our study showed requiring supplemental oxygen therapy among patients taking favipiravir was less than those taking control drugs. Dhan Bahadur Shrestha et al. also showed that patients receiving favipiravir had less need for oxygen and non-invasive mechanical ventilation ^40^.

The results of the present study showed that the groups treated with favipiravir had a lower chance of side effects compared to the control groups. This finding is consistent with the meta-analysis carried out by Shrestha et al. ^40^. Khamis et al. also found intervention with favipiravir had no significant side effects, including hyperuricemia, abnormalities in liver enzymes, or QTc prolongation ^42^.

Erdem et al. found that side effects occurred in 13% of patients during treatment with favipiravir. The most common side effects were elevation of liver enzymes, total bilirubin, and uric acid, as well as gastrointestinal disorders. This trial consists of five patients, and All five experienced mild to moderate rise in liver enzymes, three of them nausea, and one of them neutropenia. All side effects were self-limited. There was no association between underlying disease and serious side effects, and no patients stopped favipiravir due to side effects ^37^.Victoria Pilkington et al. demonstrated that patients who took favipiravir had no serious side effects. However, an increase in serum uric acid remains a concern, and the analysis of studies showed some evidence of a dose-dependent increase in this biochemical parameter. Other complications, including teratogenic potential and QTc prolongation, have not been sufficiently studied^43^. Denis Malvy et al. also reported that favipiravir is well tolerated and safe in short-term administration. However, more evidence is necessary to conclude long-term safety ^44^. Udwadia et al. reported most of the side effects were mild to moderate, and the most common side effects were an asymptomatic transient rise in serum uric acid and liver enzymes. On the other hand, gastrointestinal disorders were minimal ^39^.

Totally, intervention with favipiravir exerted minor tolerable side effects, including nausea, vomiting, diarrhea, and elevated serum transaminases. There were no serious life-threatening side effects after treatments with favipiravir. Possible side effects could not be attributed to the only consumption of favipiravir. Patients in the favipiravir groups received other drugs in all three trials ^40^.

Our analysis showed the need for admission in ICU is not statistically significant between the favipiravir groups and control groups. Khamis et al. also revealed there was no significant difference between favipiravir and hydroxychloroquine group in the case of transfer to ICU^42^ Additionally, Yan Lou et al. found only two of the 22 patients in favipiravir and one patient in baloxavir marboxil group transferred to the ICU within seven days of starting intervention ^41^.

Based on the results of the analysis, there has been a decrease in all-cause mortality in patients who took favipiravir compared to those who took control of drugs. In a study carried out by Dabbous, one patient in the hydroxychloroquine group expired. However, no death was not reported in the favipiravir group ^30^.

Considering the importance of treating patients with COVID-19, further studies on the role of favipiravir in the management of COVID-19 patients are recommended. Despite the limitation, the present study provided the information needed for treating COVID-19, suggesting that favipiravir is associated with significant clinical and laboratory improvement in most patients and it is a safe drug with no serious side effects ^37^.

There is some evidence to support the safety and tolerability of favipiravir in short-term administration. However, more evidence is necessary to evaluate the exact long-term effects of this intervention. Due to limited evidence and other specific safety concerns, caution should be considered in the widespread use of favipiravir against the COVI D-19 epidemic ^45^.

## Limitations

There are some limitations to the included studies. First, the sample size is low in each study. Second, due to multiple drug pharmacotherapy of patients with COVID-19 in the most included study, there was, therefore, a risk of influencing the efficacy and also the safety of intervention with favipiravir. Third, the dosage and duration of intervention with favipiravir are different among the included studies. Fourth, it is difficult to determine the clinical improvement found in patients treated with favipiravir from different disease severity, ages, and medical conditions in the different studies.

## Conclusion

Overall, favipiravir possibly exerted no significant beneficial effect in the term of mortality in the general group of patients with mild to moderate COVID-19. We should consider that perhaps the use of antivirals once the patient has symptoms is too late and this would explain their low efficacy in the clinical setting. There upon, more clinical trials with a larger sample size are necessary to evaluate the exact efficacy and safety of this intervention.

## Data Availability

All relevant data included in the manuscript and also is available by contacting the corresponding author.

## Authorship contribution statement

MA-Z: conceptualization, methodology, formal Analysis, writing, validation, software, project administration, supervision; SH: methodology, formal Analysis, writing, validation, software; BA: methodology, data curation, writing; FH: methodology, data curation, writing; MF: methodology, data curation, writing; RM: supervision, writing, review and editing. All authors approve the manuscript before submission.

## Funding

None

## Declaration of Competing Interest

MSN produce favipiravir (Favilow®). The authors declared no conflict of interest.

## Notes

### Competing Interest Statement

MSN produces favipiravir (Favilow). The authors declared no conflict of interest.

### Clinical Protocols

https://bmjopen.bmj.com/content/10/7/e039730

### Author Declarations

We published the protocol in the BMJ open journal. due to the design of this study, there is no need for ethics approval.

