## Supplementary material for "The efficacy and safety of Favipiravir in treatment of COVID-19: A systematic review and meta-analysis of clinical trials": Supplementary file 1.pdf

**Supplement 1: Proposed Search Strategy for PubMed**

| Search | Search strategy |
|---|---|
| **#1** | Search (("2019 nCoV" OR "2019 nCoV" OR "2019 novel coronavirus" OR "COVID-19" [Supplementary Concept] OR "severe acute respiratory syndrome coronavirus 2" [Supplementary Concept] OR "new coronavirus" OR "novel coronavirus" OR "SARS CoV-2" OR (Wuhan AND coronavirus) OR "SARS-CoV" OR "2019-nCoV" OR "SARS-CoV-2")) |
| **#2** | (favipiravir [Supplementary Concept] OR Avigan OR T-705 cpd) |
| **#3** | #1 AND #2 |
