## Supplementary material for "The efficacy and safety of Favipiravir in treatment of COVID-19: A systematic review and meta-analysis of clinical trials": Supplementary file 2.pdf

| | Random sequence generation (selection bias) | Allocation concealment (selection bias) | Blinding of participants and personnel (performance bias) | Blinding of outcome assessment (detection bias) | Incomplete outcome data (attrition bias) | Selective reporting (reporting bias) | Other bias |
|---|---|---|---|---|---|---|---|
| Cai et al/ 2020 | − | − | − | + | + | + | + |
| Chen et al/ 2020 | + | − | − | − | + | + | + |
| Dabbous et al/ 2020 | + | + | − | − | + | + | + |
| Doi et al/ 2020 | + | + | − | − | + | + | + |
| Ivashchenko et al/ 2020 | + | + | − | − | + | ? | + |
| Khamis et al/ 2021 | + | ? | − | − | + | + | + |
| Lou et al/ 2020 | + | + | − | − | + | + | + |
| Udwadia et al/ 2020 | + | + | − | − | + | + | + |
| Zhao et al/ 2021 | + | + | − | − | + | + | + |
