## Supplementary figures and images for "The efficacy and safety of Favipiravir in treatment of COVID-19: A systematic review and meta-analysis of clinical trials"

### supplementary file 3.pdf

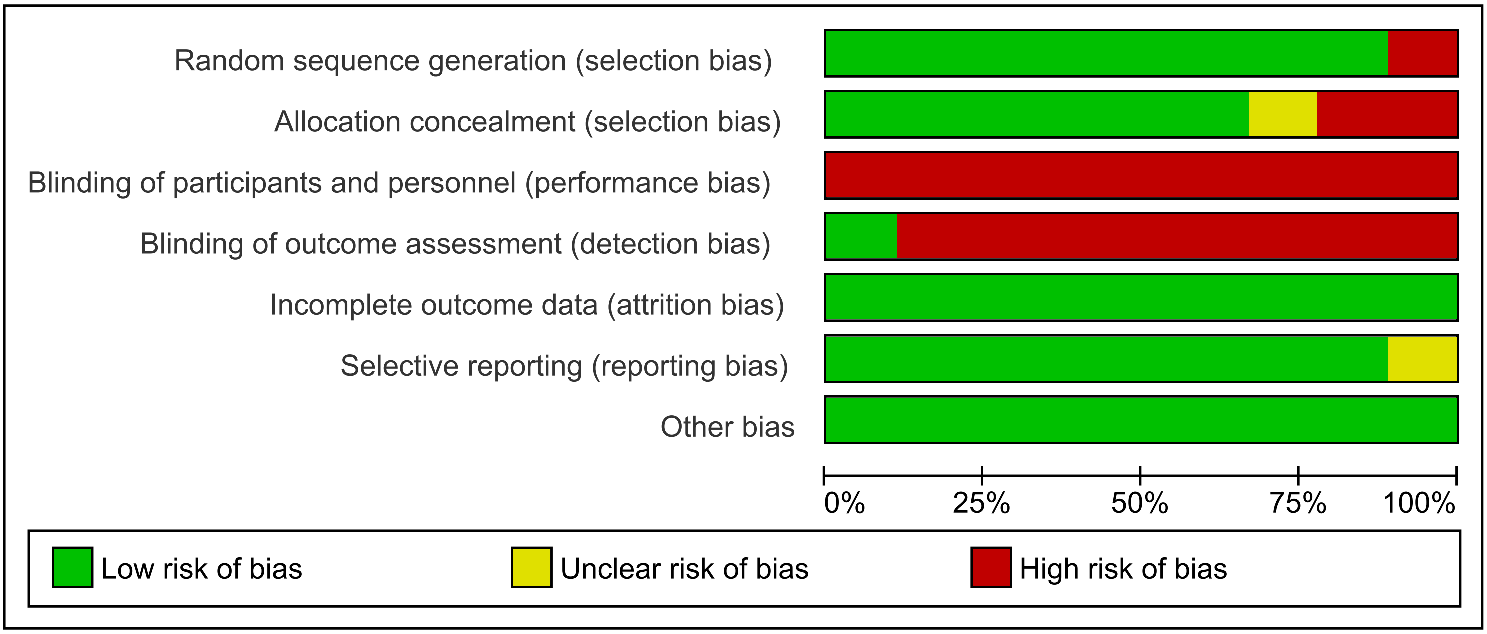
